# Toward Localizing Psychosis in Pathologically Confirmed Neurodegenerative Disease

**DOI:** 10.1101/2025.02.06.25321537

**Authors:** Andrew G. Breithaupt, Georges Naasan, Suzanne M Shdo, Lucia Lopez, Salvatore Spina, Lea Grinberg, William W Seeley, Maria Luisa Gorno-Tempini, Gil Rabinovici, Joel H. Kramer, Howard J. Rosen, Bruce L Miller, Katherine P Rankin

**Affiliations:** Memory and Aging Center, Department of Neurology, 1651 4th Street, University of California, San Francisco, CA 94158; Goizueta Institute at Emory Brain Health, Department of Neurology, Emory University School of Medicine, 6 Executive Park Drive, Atlanta, GA, 30329; The Barbara and Maurice Deane Center for Wellness and Cognitive Health, Department of Neurology, Icahn School of Medicine, Mount Sinai, New York, NY, USA

## Abstract

Psychosis is a common symptom in neurodegenerative diseases that contributes to significant patient and caregiver burden. While neuroimaging studies have implicated various brain regions in psychosis, findings have been inconsistent across different disease entities and psychosis subtypes. This study aimed to identify structural neuroanatomic changes associated with specific patterns of psychotic content across pathologically confirmed neurodegenerative diseases. We examined 283 autopsy-confirmed neurodegenerative disease cases (70 with psychosis) at a tertiary medical center, representing diverse clinical syndromes and pathologies. Psychotic content was systematically classified using standardized criteria. Voxel-based morphometry analyses of MRI were conducted to identify structural correlates of psychotic features across all syndromes, within specific clinical syndromes, and within pathological subtypes. Overall, delusions were associated with atrophy in the right temporal lobe and bilateral frontal lobes, particularly when the delusions were persecutory or paranoid. Together, these regions support processing of external stimuli, reward, emotion, self-awareness, and executive function. By contrast, misidentification delusions correlated with right ventral temporal-occipital atrophy, implicating selective disruption of ventral visual stream processing. No consistent patterns of atrophy were found with hallucinations. Our findings suggest that damage to temporal and frontal subregions predisposes individuals with neurodegeneration to develop delusions across clinical syndromes and pathologies. This study provides support for the theory that dysfunction in brain circuits supporting reward, emotion, self-awareness, processing of external sensory signals, and executive functioning can lead to new-onset delusional beliefs in neurodegenerative disease. These insights suggest shared mechanisms between “neurologic” and “psychiatric” disorders that could inform future prognostic and therapeutic approaches.

## Introduction

Psychosis is common in neurodegenerative disease and may occur in the early stages of illnes.^1–4^ Estimates of the prevalence of psychosis range from 22 to 54% in Alzheimer’s disease (AD) and from 18 to 42% in frontotemporal dementia (FTD).^5^ Psychosis increases stress, anxiety, and depression, placing a significant burden both on people suffering from the disorder, their caregivers and their health care providers.^6^ Further, , psychosis in persons with dementia is associated with earlier institutionalization and a higher cost of care making it an important target for new approaches to understanding its etiology and treatment.^7^ One feature of psychosis associated with neurodegenerative diseases is that there is often a visible anatomic substrate that may help to better understand mechanisms in psychosis with primary psychiatric disorders like schizophrenia where anatomic mechanisms are more poorly understood. There are diverse underlying biological and physiological substrates associated with different presentations of psychosis. For example, individuals with psychiatric disease, neurodegenerative disease, and eye disease have significantly different patterns of hallucinations. Likewise, rates and patterns of delusions differ among individuals with bvFTD with C9orf72 expansions, AD, and Lewy body disease (LBD)^6,8–11^. The anatomic specificity for the clinical phenomenology in psychosis requires further study.

Most research investigating the neuroimaging correlates of psychosis has been in schizophrenia, and studies in neurodegenerative disease have been comparatively scant. Still, results with both types of disorders have implicated the frontal lobe, with more varied correlates in the temporal, occipital, and parietal lobes, as well as the basal ganglia and the cerebellum^12–18^. There is relatively more neuroimaging research involving individuals with AD compared with other neurodegenerative diseases, and this work has associated psychosis with right-sided frontal and temporal atrophy or dysfunction with SPECT and PET studies^13^. Dividing studies by psychosis subtypes yields some anatomic specificity. For instance, two studies in AD found right hippocampal atrophy associated with delusions^19^, and one of these studies also found bilateral lateral temporal lobe atrophy^20^. In a study using SPECT in patients with clinically diagnosed LBD, hallucinations corresponded to hypoperfusion of the parietal and occipital association cortices, but delusional misidentifications were related to hypoperfusion of the limbic-paralimbic structures. Delusions of theft and persecution, conversely, were related to *hyper*perfusion of the frontal cortices^9^. In studies of clinically diagnosed Parkinson’s Disease (PD), hallucinations have been associated with wide ranging areas of atrophy in the frontal, parietal, hippocampal, limbic, and occipital lobes^16^, though without pathological confirmation it is unclear whether a subset of these participants actually had concomitant Lewy body disease or AD. This variability suggests that other factors like neurotransmitter changes may contribute to hallucinations in these disorders.^21^ Indeed, a recent systematic review on delusions across schizophrenia, bipolar disorder, and Alzheimer’s disease found delusions were most commonly associated with gray matter reductions in the dorsolateral prefrontal cortex, middle frontal gyrus, and superior temporal gyrus cortically, with additional correlations with the left claustrum, hippocampus, insula, amygdala, and thalamus. However, they found a great deal of anatomic variability was reported across disorders, and most studies did not clarify the nature of psychotic content beyond broad categories^22^.

Our group previously reported rates of psychosis across groups of pathology confirmed neurodegenerative diseases and noted that greater subgroup specificity was found when the nature of psychotic content was carefully delineated.^23^ For the current study, we wished to further clarify the structural neuroanatomic changes most likely to result in distinct patterns of psychosis in individuals with neurodegenerative disease, using precise endophenotyping of the psychotic content into detailed subcategories to maximize anatomic precision. Our primary goal in this exploratory study was to delineate for each specific type of psychotic content any brain-behavior relationships that were generalizable across neurodegenerative disease syndromes, while as a secondary goal we examined whether any patterns of regional atrophy were more likely to result in particular psychotic symptoms within clinical dementia syndrome or neuropathological group.

## Methods

### Participant selection

We screened all patients that came to autopsy at the University of California San Francisco Memory and Aging Center between 1999 and 2017 (N=830). Patients were excluded if full review of their pathology reports did not confirm the presence of a primary neurodegenerative neuropathology, operationalized as: (i) Alzheimer’s disease (AD) without Lewy body disease (LBD), defined as a primary pathology of Alzheimer’s disease meeting the ADNC criteria (Montine et al.,2012) with LBD showing a Parkinson’s disease Braak stage < 3; (ii) mixed LBD and Alzheimer’s disease (LBD/AD), defined as concomitant Alzheimer’s disease and LBD pathology where the Parkinson’s disease Braak stage was > 3; (iii) TDP pathology which included frontotemporal lobar degeneration with TDP inclusions (FTLD-TDP), including types A–C, as well as ALS-TDP and FTLD-MND; (iv) tau pathology including FTLD-tau, progressive supranuclear palsy (PSP), corticobasal degeneration (CBD), Pick’s disease, and argyrophilic grain disease (AGD); and (v) Other neurodegenerative disease, including FTLD FUS (Fused in Sarcoma), FTLD-Ubiquitin atypical, Huntington’s Disease, and Parkinson’s Disease (PD Braak 5). While these were the primary pathologies, almost every patient had additional copathologies (See supplement Tables S2 and S3 for detailed description).

Patients were further classified based on whether or not there was evidence that they had psychosis. Participants were further screened for having suitable structural MRI brain imaging for quantitative volumetric analysis (see “Neuroimaging acquisition and VBM preprocessing” below for more details). Individuals were excluded if they did not have data on their psychosis status reported on the Neuropsychiatric Inventory (NPI)^24^ or the Alzheimer’s Disease Research Center (ADRC) UDS Symptom Checklist. The final sample for this study included N=283 individuals, 70 of whom had MRI and psychosis, for whom comprehensive chart reviews of psychosis history were conducted, and N=213 who had MRI and no psychosis.

The neurodegenerative disease syndrome of each participant was derived by a team of expert neurologists, neuropsychologists, nurses, and other specialists as part of the patient’s involvement in a large multidisciplinary longitudinal study at our center. Primary syndrome categories observed in this sample included: Alzheimer’s disease syndrome (AD) diagnosed according to consensus clinical criteria^25^ (N=79), behavioral variant frontotemporal dementia (bvFTD; N=62),^26^ progressive supranuclear palsy syndrome (PSPS; N=38)^27^, corticobasal syndrome (CBS; N=29)^28^, amyotrophic lateral sclerosis (ALS; N=13)^29^, FTD/ALS (N=20), LBD/PD (N=4)^30,31^, and primary progressive aphasia of the semantic (N=19) and nonfluent (N=19) variants^32^ (see Table 1 for syndrome frequencies and clinical characteristics). All participants consented to have their data used for secondary analysis, and the protocol for this study was approved through the institution’s internal review board, in accordance with the Declaration of Helsinki.

**Table 1:** Patient Characteristics.

|  | Psychosis | No Psychosis | Total | P-value |
| --- | --- | --- | --- | --- |
| **Age At MRI Scan** | 64.6 ($\pm 9.2$) | 64.8 ($\pm 9.7$) | 64.8 ($\pm 9.6$) | n.s. |
| **Sex** |  |  |  | n.s. |
| Male | 38 (23.5%) | 124 (76.5%) | 162 (57.2%) |  |
| Female | 32 (26.4%) | 89 (73.6%) | 121 (42.8%) |  |
| **Clinical Dementia Rating Global Score** | | | | $< 0.0001$ |
| Mean (SD) | 1.4 ($\pm 0.7$) | 0.8 ($\pm 0.6$) | 1.0 ($\pm 0.7$) | |
| **Clinical Dementia Rating Global Score (with NACC FTLD)** | | | | $< 0.0001$ |
| Mean (SD) | 2.0 ($\pm 0.7$) | 1.2 ($\pm 0.7$) | 1.4 ($\pm 0.8$) | |
| **MMSE** | | | | $< 0.0001$ |
| Mean (SD) | 19.4 ($\pm 7.8$) | 23.2 ($\pm 6.7$) | 22.3 ($\pm 7.2$) | |
| **Clinical Diagnosis** |  |  |  |  |
| bvFTD | 24 (38.7%) | 38 (61.3%) | 62 (21.9%) |  |
| AD | 21 (26.6%) | 58 (73.4%) | 79 (27.9%) |  |
| PSP | 8 (21.1%) | 30 (78.9%) | 38 (13.4%) |  |
| CBS | 5 (17.2%) | 24 (82.8%) | 29 (10.2%) |  |
| ALS/bvFTD | 5 (25.0%) | 15 (75.0%) | 20 (7.1%) |  |
| DLB/PD | 2 (50.0%) | 2 (50.0%) | 4 (1.4%) |  |
| svPPA | 4 (21.1%) | 15 (78.9%) | 19 (6.7%) |  |
| nfvPPA | 0 (0.0%) | 19 (100.0%) | 19 (6.7%) |  |
| ALS | 1 (7.7%) | 12 (92.3%) | 13 (4.6%) |  |
| **Pathology Category** |  |  |  |  |
| TDP | 23 (29.9%) | 54 (70.1%) | 77 (27.2%) |  |
| tauopathy | 22 (19.1%) | 93 (80.9%) | 115 (40.6%) |  |
| AD | 15 (23.8%) | 48 (76.2%) | 63 (22.3%) |  |
| LBD_AD | 9 (40.9%) | 13 (59.1%) | 22 (7.8%) |  |
| Other | 1 (16.7%) | 5 (83.3%) | 6 (2.1%) |  |

### Chart Review

For the 70 individuals found to have psychosis, medical records were systematically reviewed for information about patients’ psychosis by trained coders who were blind to neuropathological diagnosis. To maximize reliability among coders, 20% of the charts of patients with psychosis were rated by two coders and discrepancies were discussed, resolved, and incorporated into an enhanced coding manual. Hallucinations were classified by sensory modality (visual, auditory, tactile, and olfactory) and by content of the hallucinated stimulus (visual misperceptions, simple visual hallucinations of shapes and colors, well-formed visual hallucinations including of people or animals, and non-visual hallucinations). Delusion subtypes were selected according to the DSM-5 criteria. Delusions were divided into five major groups including: (i) erotomanic; (ii) grandiose; (iii) jealous; (iv) persecutory; and (v) somatic (American Psychiatric Association, 2013). In addition, we added a category to capture delusions of misidentification of people, including the Capgras delusion, and for delusions regarding a particular place, including a person’s own home and reduplicative paramnesia. We further subclassified persecutory delusions into delusions of theft, intention to hurt, intrusion into home, and suspiciousness, for paranoid delusions that did not meet any of the other classifications. Finally, we included the feeling of presence, an overwhelming feeling that another person is present in the same room when no one is there, as its own category as its classification is unclear^10^. See Naasan 2021^23^ for more detailed operational definitions.

### Measures of function

Data collected at the same time as the MRI scan was used to represent each patient’s level of cognitive and behavioral functioning.

The CDR® Dementia Staging Instrument is a semi-structured interview used to characterize the cognitive/functional levels and severity of patients with dementia by rating the following six domains: memory, orientation, judgment and problem solving, community affairs, home and hobbies, and personal care ^33,34^. The CDR® Dementia Staging Instrument PLUS National Alzheimer’s Coordinating Center Behavior and Language Domains (CDR plus NACC FTLD) helps characterize the severity of FTLD-related dementia by rating two additional domains: behavior and language (Miyagawa et al., 2020). These domains are rated from normal (score of 0), minimally impaired (score of 0.5), mildly impaired (score of 1), moderately impaired (score of 2) and severely impaired (score of 3). To assess disease severity, the global CDR® and the global CDR® plus NACC FTLD scores were calculated using published scoring rules that assign weights to domains to ensure score optimization^33,35–37^. The CDR® plus NACC FTLD Sum of Boxes was calculated using the total sum of the domains to facilitate equal evaluation of all domains.

The MMSE is a brief screening test that uses dimensions such as orientation, attention, recall and language to assess severity of cognitive impairment. It is a highly verbal screening tool and is mainly sensitive to picking up lower to moderate levels of impairment^38^.

### Neuroimaging acquisition and VBM preprocessing

Participants underwent MRI brain imaging with a 3T, 1.5T, or 4T ADNI-compliant structural sequence; the acquisition details are described elsewhere^39^. If the patient already had psychosis upon initial clinic presentation (e.g., if they had begun to experience psychotic symptoms 2 years before first clinic visit), then we chose the earliest available MRI closest to that initial visit for analysis. If psychosis onset occurred after their initial clinic visit (e.g. 3 years after initial evaluation), we selected a scan within 180 days of the established psychosis onset date. For individuals with no reported psychosis, then the earliest available MRI scan was always chosen.

Structural T1-weighted images were preprocessed using SPM12. The images were visually inspected for artifacts, and underwent bias-correction, segmentation into tissue compartments, and spatial normalization using a single generative model with the standard SPM12 parameters. The default tissue probability maps for grey matter, white matter, cerebrospinal fluid, and all other voxels from SPM12 (TPM.nii) were used^40^. To optimize intersubject registration, each participant’s image was warped to a template derived from 300 confirmed neurologically healthy older adults (ages 44-86, M±SD: 67.2±7.3; 113 males, 186 females) scanned with one of three magnet strengths (1.5T, 3T, 4T), using affine and nonlinear transformations with the help of the diffeomorphic anatomical registration through exponentiated lie algebra (DARTEL) method, with standard implementation in SPM12^40^. In all preprocessing steps, default parameters of the SPM12 toolbox were used. Total volume of each tissue compartment was calculated by applying the modulated, warped and segmented masks for gray matter, white matter, and CSF to the corresponding MWS probability map for that individual, and the total intracranial volume (TIV) was derived by summing the three volumes. The spatially normalized, segmented, and modulated gray matter images were smoothed with an 8-mm FWHM isotropic Gaussian kernel for use in voxel-based morphometry analysis^40,41^.

### Statistical analysis

Patient characteristics and group frequencies were analyzed using R, and brain-behavior analyses were performed using SPM12. All VBM analyses included age, gender, magnet strength, total intracranial volume (TIV), and MMSE as nuisance covariates. <u>All-syndrome analysis:</u> First, a series of VBM analyses were performed with all N=283 individuals in the study to determine if the presence or absence of any major form of psychosis was predicted by gray matter atrophy at the voxel level, using a one-tailed t-contrast. Only results surviving a family-wise-error threshold of pFWE < 0.05 with a cluster size > 25 mm3 were considered significant. <u>Within-syndrome subgroup analyses:</u> Next, any form of psychosis that showed generalized brain-behavior relationships in the whole group that were significant at an FWE-corrected level was further examined via a series of subgroup analyses limited to a single clinical syndrome. For each syndrome group, individuals positive for that psychotic feature were compared to those without that feature. The rationale for this approach was that 1) distinct syndrome-specific patterns of neurodegeneration may cause the same psychotic symptoms through damage to different brain regions, and 2) the all-syndrome analysis could be biased if disproportionate numbers of individuals with any one clinical syndrome were represented in the psychosis group compared to the no-psychosis group, but this bias would not occur in a within-syndrome analysis. Given that this analysis was based on significantly smaller sample sizes and thus would be underpowered to detect real differences, uncorrected results at a p<0.0001 level were also considered significant. For clinical syndromes without significant results, we performed a qualitative evaluation by visually comparing the atrophy maps for each psychosis-by-syndrome subgroup to healthy controls identified with 1:1 matching based on age. <u>Within-pathology</u> <u>subgroup analyses:</u> Finally, any form of psychosis found to have significant brain correlates in the all-syndrome analysis was further examined by subgrouping individuals within primary pathology category (AD, AD/LBD, TDP, Tau) and comparing those with and without the particular psychotic symptom. As with the within-syndrome analyses, small sample sizes made these analyses somewhat underpowered, and thus uncorrected results at p<0.0001 were accepted. This analysis was performed to evaluate the hypothesis that patients with more homogenous neuropathology might show similar psychotic symptoms resulting from pathology-specific patterns of focal damage. This hypothesis was based on the idea that neuropathologies might be considered a “biotype”, direct analysis of which has been advocated by the psychiatry research community for over at least a decade in an attempt to deprioritize DSM and ICD clinical diagnoses, which are “taxed by high degrees of inter-class overlaps, comorbidity and heterogeneity, as well as diverse disease course and response to treatment”^42^.

Family-wise error correction was used to identify the lower bound of the significance threshold for the VBM analysis. In this correction, the maximum t-values of the imaging data compared to behavioral data were re-sampled using the Monte Carlo approach, which includes 1000 permutations of the error distribution. This was used to create a custom map of the error distribution based on the data set, i.e., distribution of maximum t-values when no true relationship exists. The t-value at the 95th percentile of this distribution was taken as the custom cut-off threshold, rendering t-values on or above this cut-off significant at a family-wise error corrected level of p < .05^43^.

## Results

The demographic characteristics of all participants, as well as their clinical syndrome and pathologic diagnosis groupings by psychosis status, are presented below in Table 1. There were no statistical differences in gender distribution or age at MRI scan across clinical syndrome groups. There was a statistically significant difference in level of function between patients who did and did not have psychosis, with worse scores in the psychosis group on both standard and NACC FTLD versions of the CDR Global score, and the MMSE total score. Of note, however, on average both psychosis and non-psychosis groups in our sample were very early in their disease process at the analyzed visit, averaging a CDR global score of M=1/SD=0.7, which is consistent with “very mild dementia”, thus this statistical difference may not signify a clinically meaningful discrepancy.

### All-syndrome analyses

Some forms of psychosis that were part of the chart review appeared in few to no patients in the sample, and thus could not be analyzed (less than 5). The forms of delusions analyzed included grandiose delusions, paranoid delusions, misidentification delusions, delusions regarding a particular place, and persecutory delusions, including its subcategories of intention to hurt, theft, and suspiciousness. The forms of hallucinations analyzed included visual misperceptions, formed hallucinations (including of people, animals, insects), hallucinations of shapes, and hallucinations of voices. All analyzed psychosis forms were evaluated with whole-brain VBM using all patients regardless of clinical syndrome or pathologic diagnosis. The forms of psychosis with significant correlations (pFWE<0.05) between the psychosis and brain volume are presented below (Table 2, Figure 1).

**Figure 1:**
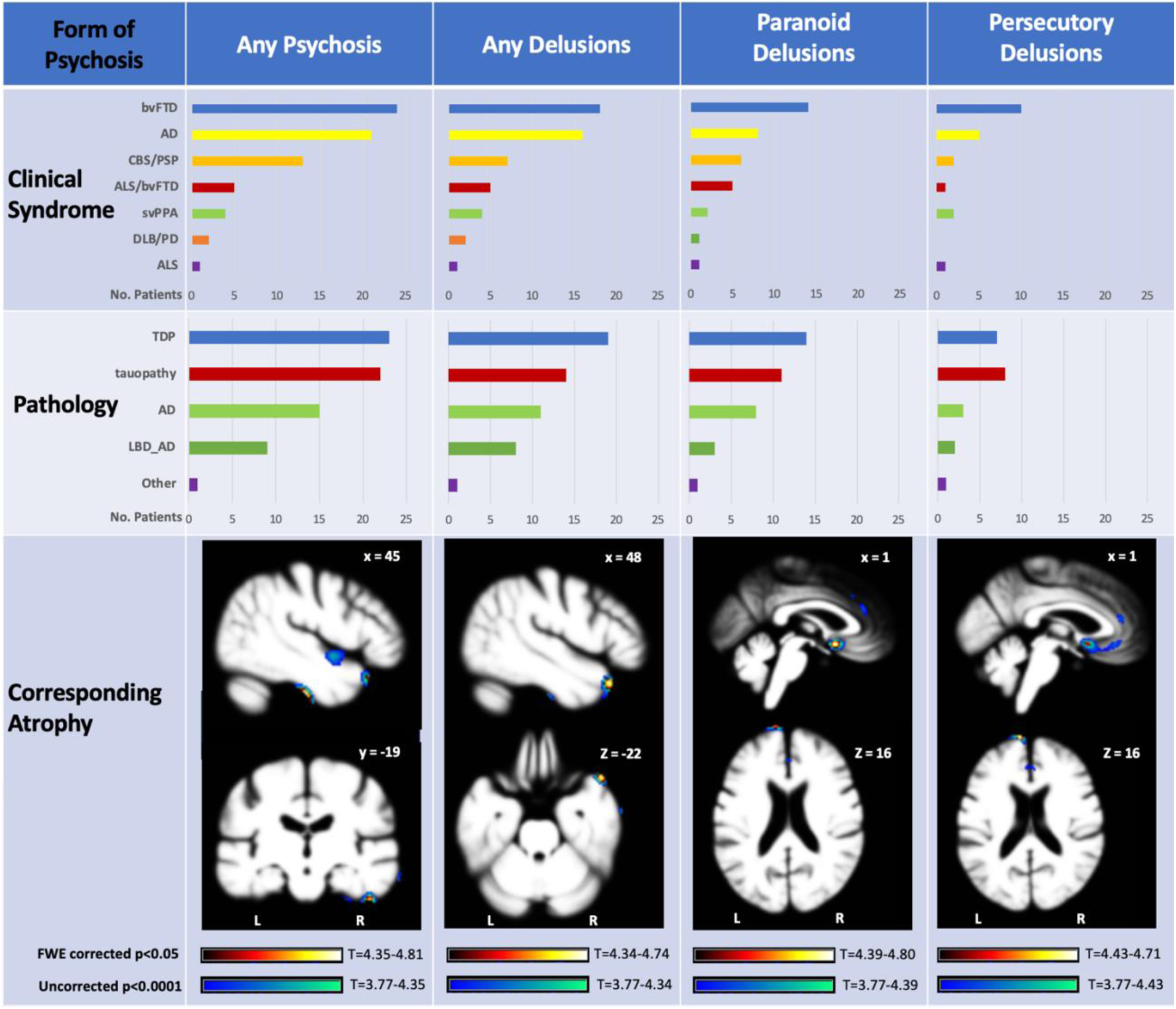
Areas of significant atrophy by form of psychosis

**Table 2.**
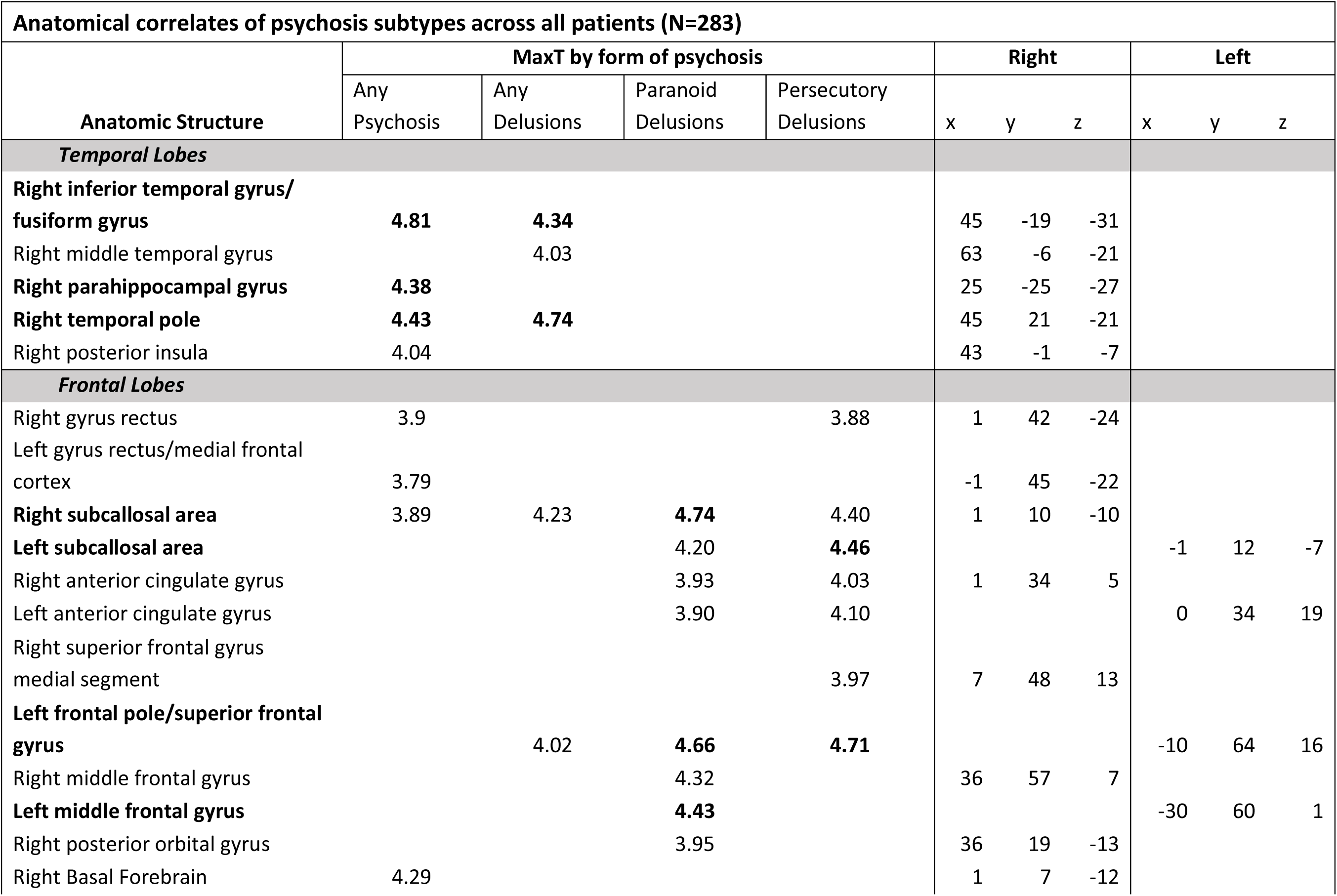
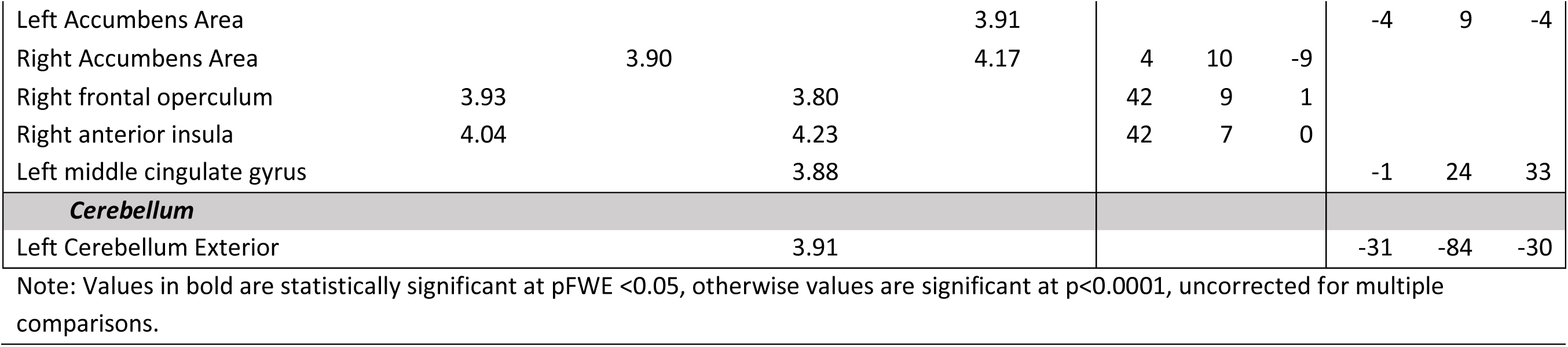

### Any psychosis

The psychosis group (N=70) had significantly greater atrophy than the non-psychosis group (N=213) in the right inferior and anterior temporal lobe, specifically the temporal pole, parahippocampal gyrus, and a contiguous area of atrophy overlapping with the inferior temporal gyrus and fusiform gyrus (pFWE<0.05). Prominent areas of atrophy that did not survive family-wise error correction but were significant at a p<0.0001 level were found in the frontal lobes as well, including the bilateral gyrus rectus, medial frontal cortex, right subcallosal area, right basal forebrain, right frontal operculum, and right anterior insula.

### Any delusions

Patients with delusions (N=53) had two areas where they showed significantly greater atrophy than those without delusions (N=230). These strongly overlapped with the regions described above found for “any psychosis”, including the right temporal pole, and the right inferior temporal gyrus (pFWE<0.05), as well as similar right frontal and middle temporal regions that did not survive FWE correction that were listed above (subcallosal area and accumbens, left frontal pole, superior frontal gyrus medial segment, middle temporal gyrus).

### Paranoid delusions

In individuals with paranoid delusions (N=37), the frontal lobe atrophy patterns seen at an uncorrected threshold in the “any delusions” analysis (above) became significantly worse than patients without paranoid delusions (N=246) (pFWE<0.05). This included the right subcallosal area, a contiguous area of atrophy overlapping the left frontal pole and superior frontal gyrus, as well as the left middle frontal gyrus. Regions not surviving FWE correction included the left subcallosal area, bilateral anterior cingulate gyri, right middle frontal gyrus, left middle cingulate gyrus, and the left cerebellum exterior.

### Persecutory delusions

In individuals with delusions that were specifically classified as persecutory (N=21), the areas of atrophy previously found in individuals with paranoid delusions were again prominent, including the subcallosal area and left anterior frontal lobe. Both the left and right subcallosal areas appeared to predict persecutory delusions, although in this analysis the left subcallosal area survived FWE correction while the right subcallosal area did not by a marginal degree, as can be seen in Table 2 and Figure 1. Areas of atrophy that did not survive FWE correction but were significant at p<0.0001 included the bilateral accumbens, bilateral anterior cingulate, and right gyrus rectus.

### Delusions of misidentification

The atrophy pattern specific to individuals with delusions of misidentification (N=11) was very close to the FWE-corrected threshold, but dropped below this threshold when MMSE was added as a covariate. We report these results because potentially they are meaningful but could not survive our conservative statistical threshold. Two regions of atrophy significant at an uncorrected p of <0.0005 were in the ventral visual pathway, including the right posterior inferior temporal gyrus and the right posterior middle temporal gyrus.

### Additional subtypes of delusions

No other subtype of delusions showed significant differences between individuals with and without that symptom, including grandiose delusions and delusions regarding a particular place. Two subcategories of persecutory delusions were significant at p<0.0001: intention to hurt (n=6) and delusions of suspiciousness (n=13). Intention to hurt revealed a small area of atrophy at the right entorhinal area, which did not overlap with previous areas of significance with any analysis. Delusions of suspiciousness revealed atrophy in the right superior frontal gyrus medial segment and the right middle frontal gyrus.

### Any hallucinations

Though there was a relatively large sample size of patients with hallucinations (N=39), when their patterns of atrophy were compared to those without, no areas of associated atrophy were found, even at an uncorrected p-value threshold of p<0.0005.

### Additional subtypes of hallucinations

No other subtype of hallucination showed significant differences in atrophy pattern between individuals with and without that symptom, including visual misperceptions, formed hallucinations (including of people, animals, insects), hallucinations of shapes, and hallucinations of voices.

### Within-syndrome subgroup analyses

In the PSP-only subgroup analysis, individuals with PSP and any psychosis showed significantly greater atrophy in the right temporal pole (pFWE<0.05) than those with PSP and no psychosis. In the AD group, individuals with any psychosis showed significantly greater atrophy (uncorrected p< 0.0001) in the right fusiform gyrus, right hippocampal gyrus, and right temporal pole. svPPA patients with persecutory delusions showed significantly greater atrophy of the left superior frontal gyrus and middle frontal gyrus (however, of note, this was a small sample size of 2 with persecutory delusions versus 17 without). Neither the bvFTD or CBS syndrome groups showed any significant within-syndrome differences in atrophy between the subgroup with and without psychosis. The other diagnostic syndrome groups were considered too small, or too unbalanced, to perform within-group comparisons of individuals with and without psychosis, including the ALS, nfvPPA, and DLB/PD syndrome groups.

### Within-pathology subgroup analyses

Patients were then divided into four major groups based on their primary neuropathology, as in Naasan 2023, with the caveat that almost all patients had additional secondary pathologies, some of which were significant and likely contributed to their overall clinical syndrome. Details on the presence of copathologies are found in the Supplement (Tables S2 and S3). A subgroup analysis was performed within each major pathology group for each form of psychosis that was found to be significant after FWE correction in the all-patient analysis described above. See supplementary Figure S1 for an overview of significant areas of atrophy (pFWE<0.05) and supplementary Table S4 for all areas of significant atrophy.

### Primary TDP pathology group (N=77)

Within the TDP pathology group, individuals with any psychosis (N=23) showed significantly greater atrophy in the right superior frontal gyrus (uncorrected p<0.0001) compared to those without psychosis (N=54). Also, those with persecutory delusions (N=7) showed significantly greater atrophy in the left superior frontal gyrus compared to those with TDP pathology and no persecutory delusions.

### Primary tau pathology group (N=115)

In the subset of patients with tau pathology, individuals with any psychosis (N=22) showed significantly greater atrophy mainly in the right > left frontal lobe, including the right superior frontal gyrus medial segment, with an additional area in the right temporal pole, all of which survived FWE correction. Individuals with any psychosis also showed significantly greater atrophy in the right entorhinal area, right middle frontal gyrus, right subcallosal area, and the left gyrus rectus (uncorrected p<0.0001). Patients with paranoid delusions (N=14) also had significantly greater atrophy in the right temporal pole, right superior frontal gyrus, and the right subcallosal area (uncorrected p<0.0001). Patients with paranoid delusions additionally had significantly greater atrophy in the left anterior orbital gyrus and left subcallosal area (uncorrected p<0.0001). Similar to the “any delusions” analysis, patients with paranoid delusions (N=11) had significantly greater atrophy in the right temporal pole, right superior frontal gyrus medial segment, and bilateral subcallosal areas, but additionally revealed atrophy in the right medial frontal cortex, the right gyrus rectus, the pregenual area of the bilateral anterior cingulate gyrus, and the left frontal pole (uncorrected p<0.0001). Patients with persecutory delusions (N=8) had significantly greater atrophy in the R>L medial frontal lobes, particularly in the bilateral subcallosal area, the right gyrus rectus, superior frontal gyrus medial segment, as well as the anterior frontal lobes including the right and left frontal poles, all of which survived FWE correction. In addition, patients with persecutory delusions had significantly greater atrophy in the left superior frontal gyrus medial segment, left gyrus rectus, left medial orbital gyrus, right medial and middle frontal gyrus, right posterior orbital gyrus, anterior subgenual area of the right anterior cingulate gyrus, bilateral accumbens areas, and the right hippocampus (uncorrected p<0.0001).

### Primary AD pathology group (N=63)

Analysis of AD patients with any psychosis (N=15) and any delusions (N=11) found that they had significantly greater atrophy in the superior occipital gyrus, which survived FWE correction. AD patients with any psychosis also showed significantly greater atrophy in the right angular gyrus, right fusiform gyrus, and right parahippocampal gyrus (uncorrected p<0.0001). Finally, AD patients with delusions of misidentification (N=3) had significantly greater atrophy in the right middle occipital gyrus than AD patients without this form of psychosis (uncorrected p<0.0001).

### Primary DLB/AD pathology group (N=22)

In the mixed LBD and AD pathology group, there were no significant differences in atrophy between patients with any of the forms of psychosis when they were compared to those without that symptom.

## Discussion

This study examined a large group of individuals with neuropathologically-confirmed neurodegenerative disease across a wide range of dementia syndromes, and identified a number of generalizable brain-behavior relationships where areas of atrophy were associated with forms of psychosis. None of these subjects suffered from psychosis before the onset of their neurodegenerative disorders suggesting that the regional MRI changes associated with psychosis were pathogenic. In our sample, delusions, and particularly persecutory or paranoid delusions, were most consistently associated with atrophy in the right temporal lobe and bilateral frontal lobes. The observed brain regions are involved in processing external stimuli, reward, and emotion, as well as self-awareness and executive functioning, providing some insight into how degeneration of specific cognitive circuits may lead to delusional beliefs in these patients. Finally, misidentification delusions were found to be significantly predicted by atrophy in the right ventral temporal-occipital regions that potentially related to ventral visual stream processing and were strongly associated with AD but not FTD. No regional atrophy predicted hallucinations.

### Atrophy patterns associated with delusions

We found that atrophy in the right temporal lobe was a significant predictor of psychosis overall, and particularly of delusions. The majority of regions identified are known to perform processing and comprehension of external stimuli, including regions specific to person perception. These included the right temporal pole, which is a processing hub for both socioemotional as well as more generalized semantic information^44^, the right fusiform gyrus which supports object, building and facial recognition^45^, as well as the right inferior temporal gyrus, which has been associated with visual perceptual processing and object identification^46,47^. Another recent study by Kumfor and colleagues examining the anatomy underlying delusions in patients with clinically diagnosed neurodegenerative disease also found temporal pole and inferior temporal gyrus atrophy, though their results showed strong correlates on the left-side^48^.

When delusion subtypes were examined more closely, paranoid and persecutory delusions were associated with frontal and temporal regions involved in mood and autonomic regulation, reward processing, insight, and error monitoring. The atrophy pattern associated with paranoia and persecutions was especially strong in the bilateral subcallosal area of the anterior cingulate. The subcallosal area has strong connections to the limbic system and is involved in mood regulation^49^, autonomic regulation^50^, reward^51,52^, and has emerged as an area important in mood and affective disorders^53^. Previous data on the involvement of the subcallosal area in psychosis has varied, but overall it supports an association^54–56^. A relatively recent meta-analysis found that while there has been great heterogeneity in brain atrophy across patients with schizophrenia, one area of homogeneity among this group is comparatively smaller volumes in the anterior cingulate cortex^57^, though they did not subdivide the ACC thus the distinct contribution of the subcallosal area remained unclear.

Also, paranoia and persecutory delusions were predicted by significantly lower volume in the left frontal cortex, specifically areas involved with reward processing, insight, and error monitoring. These areas were predominantly in 1) the left middle frontal gyrus (specifically the dorsolateral prefrontal cortex) in paranoid delusions and 2) an area of atrophy overlapping with the left frontal pole and the medial superior frontal gyrus in both paranoid and persecutory delusions. The frontal pole functions to monitor and process the value or reward behind different options and actions^58,59^. Left frontal pole volume in patients with schizophrenia has been shown to correlate with insight, specifically the ability to observe their own mental processes and consider alternative explanations^60^. The middle frontal gyrus, also found in our study to be a region in which atrophy significantly predicted paranoia, is an area that is highly relevant for cognition and mood^61^ as well as error monitoring^62^. Our result is consistent with past studies finding decreased activity of the dorsolateral prefrontal cortex as well as decreased connectivity of this region with task-related brain regions in patients with schizophrenia, thought to result in impairments in coordinating thoughts and actions to facilitate goal-oriented behavior (i.e. cognitive control)^63^. A recent systematic review on delusions in schizophrenia, bipolar disorder, and AD found the dorsolateral prefrontal cortex was one of the areas most likely to contribute to delusions across groups^22^. A large study of patients with delusions who were clinically diagnosed with a neurodegenerative disease also found right middle frontal gyrus atrophy^48^. Finally, a past SPECT study in patients with clinically diagnosed LBD with delusions of theft and persecution reported dysfunction in areas similar to our results, including the left medial superior frontal gyrus and bilateral middle frontal gyri^9^.

Our study also found a weaker potential association between delusions of misidentification, where people believe that a person, place, or object they are seeing has been replaced, and atrophy of the posterior aspect of the right inferior and middle temporal gyri. These are areas associated with the ventral visual stream, consistent with dysfunction with processing information about face, place or object identity^64^. One study of patients with schizophrenia found volume reduction in the bilateral inferior temporal gyri and left middle temporal gyrus among patients with chronic schizophrenia^46^, which may contribute to the impairments with visual perceptual processing seen in schizophrenia^47^. In patients with clinically diagnosed AD, one study found an association between misidentification delusions and left parahippocampal volume reduction^65^. Also, in a study of clinically diagnosed LBD, which often has overlap with AD pathology, misidentification delusions were associated with hypoperfusion using SPECT in the left hippocampus, insula, inferior frontal gyrus, and ventral striatum/accumbens area^9^. The discrepancies in findings may be related to differences in patient population, or the somewhat broad definition of misidentification delusions (which included patients with hallucinations in one of the studies). Regardless, together with these studies, our results support the hypothesis that in neurodegenerative disease, disruptions to object identification may cause patients to develop misidentification delusions.

### Hallucinations

Our study did not find any areas of atrophy that corresponded with the presence of hallucinations in our patients, even at a low statistical threshold. Other studies have found wide ranging yet inconsistent results. In some studies of Parkinson’s disease, hallucinations have been associated with decreased cortical thickness in a wide range of brain regions including the right hemispheric frontal, occipital, parietotemporal, and insular regions in both hemispheres, in the basal ganglia and hippocampi, as well as the thalamus^12,16^. The supramarginal gyrus has been associated with hallucinations both in patients with clinically diagnosed AD^17^ as well as in patients with clinically diagnosed LBD^9^. As it is rare for hallucinations to present without delusions in patients with AD^13^, there may be a significant portion of patients with hallucinations in these prior studies who also have misidentification delusions driving those findings. It is notable that in our study, supramarginal gyrus atrophy was associated with misidentification delusions in our patients with AD pathology at a statistically weaker, uncorrected threshold. Although two of our three patients with misidentification delusions also had hallucinations, it is unlikely the hallucinations drove our finding given there were no areas of significance found when analyzing by hallucinations alone. Our study’s inability to detect regions of atrophy that consistently predicted hallucinations might have been due to the diversity of anatomy across all neurodegenerative syndromes in our sample, or inadequate sample size when analyzing by each pathology and clinical syndrome subgroup. Also, it is possible that hallucinations are mediated by more of a dynamic, functional, or neurochemically-mediated process and atrophy to one or more regions may not be consistently adequate or necessary to cause hallucinations in patients with neurodegeneration. We suspect that the cholinergic deficit associated with AD and DLB is a contributor to hallucinations in these patients and this chemical deficit is not detected by measurements of atrophy.

### Clinical implications for neurodegenerative disease

When analyzed by clinical syndrome subgroup, our study had much weaker statistical power to detect significant group differences or brain-behavior relationships. Only the PSP and svPPA groups had statistically significant areas of atrophy predicting subtypes of psychosis that reflected the generalizable results from the transdiagnostic analyses. In the PSP group, psychosis predicted significant atrophy in the right temporal pole. In the svPPA group those with persecutory delusions showed significantly greater atrophy of the left superior frontal gyrus and middle frontal gyrus than those without, although it should be noted this was calculated upon a particularly small sample size for the VBM group comparison. Analysis by pathology groups reiterated many of the findings in the overall sample, including associations between atrophy in right temporal and bilateral subcallosal areas with delusions in the tau pathology subgroup.

Our results suggest that damage to subregions of the right temporal and frontal lobes predispose individuals with neurodegeneration to develop delusional ideation. Localization of psychosis continues to be challenging in primary psychiatric disorders, however, the key areas found in our study with neurodegeneration overlap with findings of other studies, mostly in patients with schizophrenia. This gives a higher degree of confidence these regions do make a meaningful contribution to psychosis pathophysiology in both patients with primary psychiatric disease and dementia. Moreover, the dysfunction expected to arise from damage to the regions identified in our study reflect theoretical models of the cognitive basis of psychosis, particularly the hypothesis that dysfunctional processing and comprehension of external stimuli ^44–47,66^ lead to misinterpretation of reality and therefore can cause a patient to arrive at incorrect conclusions. This is similar to the theory of sensory processing deficits leading to psychosis in schizophrenia^67,68^. In addition, dysfunction in brain systems related to emotion and reward may cause the individual to inaccurately label the importance of different stimuli, such as feeling a sense of reward when finding a spurious connection between unrelated or neutral stimuli, which aligns with the aberrant salience theory of schizophrenia ^69,70^.

Understanding the cognitive basis of psychosis can aid in the clinic. In the more immediate term, it can help to provide key information to patients and families, unveiling the mysteries associated with these often-disturbing symptoms. In the long term, more precise information may guide the development of more tailored prognostic and therapeutic approaches. If evidence continues to suggest there could be a common pathophysiologic thread to psychotic symptoms seen in both “neurologic” and “psychiatric” disorders, it will become even more important for psychiatry and neurology to work together in their study and management of different psychosis subtypes. One remarkable aspect of our findings is that all of our subjects were psychosis free before the onset of dementia so the specific correlations in right frontal and temporal regions are more likely to be etiological for the delusions that emerged as their disorders progressed.

### Limitations and conclusions

While our study is unique for its large sample and precise characterization of psychosis symptoms in neurodegenerative disease, it also has several limitations, including its retrospective nature. In particular, there were small sample sizes for some of the delusion types, particularly within pathology groups, which may have led to a lack of power to detect significant findings within some of these psychosis subtypes. Using a lesion model made it difficult to determine if a brain structure directly mediates the psychosis subtype, or if damage to that structure results in disconnection from other regions that lead to psychosis. Also, at least in patients with AD, hallucinations have been associated with more severe dementia, whereas our patients at the time of evaluation had relatively low CDR and MMSE scores, suggesting our patients were at the beginning stages of their neurodegenerative disease^71^; therefore, it may be that if our patients’ psychosis symptoms were reassessed at more advanced stages of their dementia, a larger sample would have been found and a greater degree of atrophy significantly predicting psychosis. Overall, however, this study provides more support for the theory that dysfunction in brain circuits supporting reward, emotion, self-awareness, processing of visual signals, and executive functioning can lead to new-onset delusional beliefs in patients with neurodegenerative disease.

## Data Availability

All data produced in the present study that can be de-identified are available upon reasonable request to the authors.

## REFERENCES

1. Steinberg M, Shao H, Zandi P, et al. Point and 5-year period prevalence of neuropsychiatric symptoms in dementia: the Cache County Study. International Journal of Geriatric Psychiatry. 2008;23(2):170–177. doi:10.1002/gps.1858

2. Lyketsos CG, Sheppard JM, Steinberg M, et al. Neuropsychiatric disturbance in Alzheimer’s disease clusters into three groups: the Cache County study. Int J Geriatr Psychiatry. 2001;16(11):1043–1053. doi:10.1002/gps.448

3. Haddad PM, Benbow SM. Visual hallucinations as the presenting symptom of senile dementia. Br J Psychiatry. 1992;161:263–265. doi:10.1192/bjp.161.2.263

4. Auning E, Rongve A, Fladby T, et al. Early and presenting symptoms of dementia with lewy bodies. Dement Geriatr Cogn Disord. 2011;32(3):202–208. doi:10.1159/000333072

5. Cressot C, Vrillon A, Lilamand M, et al. Psychosis in Neurodegenerative Dementias: A Systematic Comparative Review. Journal of Alzheimer’s Disease. 2024;99(1):85–99. doi:10.3233/JAD-231363

6. Lyketsos CG, Miller DS, Neuropsychiatric Syndromes Professional Interest Area of the International Society to Advance Alzheimer’s Research and Treatment. Addressing the Alzheimer’s disease crisis through better understanding, treatment, and eventual prevention of associated neuropsychiatric syndromes. Alzheimers Dement. 2012;8(1):60–64. doi:10.1016/j.jalz.2011.11.001

7. Herrmann N, Lanctôt KL, Sambrook R, et al. The contribution of neuropsychiatric symptoms to the cost of dementia care. Int J Geriatr Psychiatry. 2006;21(10):972–976. doi:10.1002/gps.1594

8. Dudley R, Aynsworth C, Mosimann U, et al. A comparison of visual hallucinations across disorders. Psychiatry Res. 2019;272:86–92. doi:10.1016/j.psychres.2018.12.052

9. Nagahama Y, Okina T, Suzuki N, Matsuda M. Neural correlates of psychotic symptoms in dementia with Lewy bodies. Brain. 2010;133(Pt 2):557–567. doi:10.1093/brain/awp295

10. Nagahama Y, Okina T, Suzuki N, Matsuda M, Fukao K, Murai T. Classification of psychotic symptoms in dementia with Lewy bodies. Am J Geriatr Psychiatry. 2007;15(11):961–967. doi:10.1097/JGP.0b013e3180cc1fdf

11. Shinagawa S, Nakajima S, Plitman E, et al. Psychosis in frontotemporal dementia. J Alzheimers Dis. 2014;42(2):485–499. doi:10.3233/JAD-140312

12. Ffytche DH, Pereira JB, Ballard C, Chaudhuri KR, Weintraub D, Aarsland D. Risk factors for early psychosis in PD: insights from the Parkinson’s Progression Markers Initiative. J Neurol Neurosurg Psychiatry. 2017;88(4):325–331. doi:10.1136/jnnp-2016-314832

13. Ismail Z, Creese B, Aarsland D, et al. Psychosis in Alzheimer disease — mechanisms, genetics and therapeutic opportunities. Nat Rev Neurol. 2022;18(3):131–144. doi:10.1038/s41582-021-00597-3

14. Lee SE, Khazenzon AM, Trujillo AJ, et al. Altered network connectivity in frontotemporal dementia with C9orf72 hexanucleotide repeat expansion. Brain. 2014;137(Pt 11):3047–3060. doi:10.1093/brain/awu248

15. Devenney EM, Landin-Romero R, Irish M, et al. The neural correlates and clinical characteristics of psychosis in the frontotemporal dementia continuum and the C9orf72 expansion. Neuroimage Clin. 2017;13:439–445. doi:10.1016/j.nicl.2016.11.028

16. Nishio Y, Yokoi K, Uchiyama M, et al. Deconstructing psychosis and misperception symptoms in Parkinson’s disease. J Neurol Neurosurg Psychiatry. 2017;88(9):722–729. doi:10.1136/jnnp-2017-315741

17. Donovan NJ, Wadsworth LP, Lorius N, et al. Regional cortical thinning predicts worsening apathy and hallucinations across the Alzheimer’s disease spectrum. Am J Geriatr Psychiatry. 2014;22(11):1168–1179. doi:10.1016/j.jagp.2013.03.006

18. Murray PS, Kumar S, DeMichele-Sweet MAA, Sweet RA. Psychosis in Alzheimer’s Disease. Biol Psychiatry. 2014;75(7):542–552. doi:10.1016/j.biopsych.2013.08.020

19. Serra L, Perri R, Cercignani M, et al. Are the behavioral symptoms of Alzheimer’s disease directly associated with neurodegeneration? J Alzheimers Dis. 2010;21(2):627–639. doi:10.3233/JAD-2010-100048

20. Almeida FC, Jesus T, Coelho A, et al. Psychosis in Alzheimer’s disease is associated with specific changes in brain MRI volume, cognition and neuropathology. Neurobiology of Aging. 2024;138:10–18. doi:10.1016/j.neurobiolaging.2024.02.013

21. Perry EK, Perry RH. Neurochemistry of consciousness: cholinergic pathologies in the human brain. In: Progress in Brain Research. Vol 145. Acetylcholine in the Cerebral Cortex. Elsevier; 2004:287–299. doi:10.1016/S0079-6123(03)45020-6

22. Rootes-Murdy K, Goldsmith DR, Turner JA. Clinical and Structural Differences in Delusions Across Diagnoses: A Systematic Review. Front Integr Neurosci. 2022;15:726321. doi:10.3389/fnint.2021.726321

23. Naasan G, Shdo SM, Rodriguez EM, et al. Psychosis in neurodegenerative disease: differential patterns of hallucination and delusion symptoms. Brain. 2021;144(3):999–1012. doi:10.1093/brain/awaa413

24. Cummings JL, Mega M, Gray K, Rosenberg-Thompson S, Carusi DA, Gornbein J. The Neuropsychiatric Inventory: comprehensive assessment of psychopathology in dementia. Neurology. 1994;44(12):2308–2314. doi:10.1212/wnl.44.12.2308

25. McKhann GM, Knopman DS, Chertkow H, et al. The diagnosis of dementia due to Alzheimer’s disease: Recommendations from the National Institute on Aging-Alzheimer’s Association workgroups on diagnostic guidelines for Alzheimer’s disease. Alzheimers Dement. 2011;7(3):263–269. doi:10.1016/j.jalz.2011.03.005

26. Rascovsky K, Hodges JR, Knopman D, et al. Sensitivity of revised diagnostic criteria for the behavioural variant of frontotemporal dementia. Brain. 2011;134(9):2456–2477. doi:10.1093/brain/awr179

27. Hoglinger GU, Respondek G, Stamelou M, et al. Clinical Diagnosis of Progressive Supranuclear Palsy: The Movement Disorder Society Criteria. Mov Disord. 2017;32(6):853–864. doi:10.1002/mds.26987

28. Armstrong MJ, Litvan I, Lang AE, et al. Criteria for the diagnosis of corticobasal degeneration. Neurology. 2013;80(5):496–503. doi:10.1212/WNL.0b013e31827f0fd1

29. Brooks BR, Miller RG, Swash M, Munsat TL, World Federation of Neurology Research Group on Motor Neuron Diseases. El Escorial revisited: revised criteria for the diagnosis of amyotrophic lateral sclerosis. Amyotroph Lateral Scler Other Motor Neuron Disord. 2000;1(5):293–299. doi:10.1080/146608200300079536

30. McKeith IG, Boeve BF, Dickson DW, et al. Diagnosis and management of dementia with Lewy bodies: Fourth consensus report of the DLB Consortium. Neurology. 2017;89(1):88–100. doi:10.1212/WNL.0000000000004058

31. Postuma RB, Berg D, Stern M, et al. MDS clinical diagnostic criteria for Parkinson’s disease. Mov Disord. 2015;30(12):1591–1601. doi:10.1002/mds.26424

32. Gorno-Tempini ML, Hillis AE, Weintraub S, et al. Classification of primary progressive aphasia and its variants. Neurology. 2011;76(11):1006–1014. doi:10.1212/WNL.0b013e31821103e6

33. Morris JC. The Clinical Dementia Rating (CDR): current version and scoring rules. Neurology. 1993;43(11):2412–2414. doi:10.1212/wnl.43.11.2412-a

34. Hughes CP, Berg L, Danziger WL, Coben LA, Martin RL. A new clinical scale for the staging of dementia. Br J Psychiatry. 1982;140:566–572. doi:10.1192/bjp.140.6.566

35. Knopman DS, Kramer JH, Boeve BF, et al. Development of methodology for conducting clinical trials in frontotemporal lobar degeneration. Brain. 2008;131(Pt 11):2957–2968. doi:10.1093/brain/awn234

36. Knopman D, Weintraub S, Pankratz V. Language and Behavior Domains Enhance the Value of the Clinical Dementia Rating Scale. Alzheimers Dement. 2011;7(3):293–299. doi:10.1016/j.jalz.2010.12.006

37. Miyagawa T, Brushaber D, Syrjanen J, et al. Utility of the global CDR® plus NACC FTLD rating and development of scoring rules: Data from the ARTFL/LEFFTDS Consortium. Alzheimers Dement. 2020;16(1):106–117. doi:10.1002/alz.12033

38. Tombaugh TN, McIntyre NJ. The mini-mental state examination: a comprehensive review. J Am Geriatr Soc. 1992;40(9):922–935. doi:10.1111/j.1532-5415.1992.tb01992.x

39. Rijpma MG, Montembeault M, Shdo S, Kramer JH, Miller BL, Rankin KP. Semantic Knowledge of Social Interactions is Mediated by the Hedonic Evaluation System in the Brain. Cortex. 2023;161:26–37. doi:10.1016/j.cortex.2022.12.015

40. Ashburner J, Barnes G, Chen C, Deaunizeau J, Flandin G, Friston K, Kiebel S, Kilner J, Litvak V, Moran R, Penny W. SPM12 manual. Published online 2016. http://www.fil.ion.ucl.ac.uk/spm/doc/spm12manual

41. Acosta-Cabronero J, Williams GB, Pereira JMS, Pengas G, Nestor PJ. The impact of skull-stripping and radio-frequency bias correction on grey-matter segmentation for voxel-based morphometry. Neuroimage. 2008;39(4):1654–1665. doi:10.1016/j.neuroimage.2007.10.051

42. Lee TY, Jo HJ, Koike S, Raballo A. Editorial: Biotyping in Psychiatry. Frontiers in Psychiatry. 2022;13. Accessed June 14, 2022. https://www.frontiersin.org/article/10.3389/fpsyt.2022.844206

43. Kimberg DY, Coslett HB, Schwartz MF. Power in Voxel-based lesion-symptom mapping. J Cogn Neurosci. 2007;19(7):1067–1080. doi:10.1162/jocn.2007.19.7.1067

44. Córcoles-Parada M, Ubero-Martínez M, Morris RGM, Insausti R, Mishkin M, Muñoz-López M. Frontal and Insular Input to the Dorsolateral Temporal Pole in Primates: Implications for Auditory Memory. Frontiers in Neuroscience. 2019;13. Accessed June 9, 2022. https://www.frontiersin.org/article/10.3389/fnins.2019.01099

45. Weiner KS, Zilles K. The anatomical and functional specialization of the fusiform gyrus. Neuropsychologia. 2016;83:48–62. doi:10.1016/j.neuropsychologia.2015.06.033

46. Onitsuka T, Shenton ME, Salisbury DF, et al. Middle and Inferior Temporal Gyrus Gray Matter Volume Abnormalities in Chronic Schizophrenia: An MRI Study. AJP. 2004;161(9):1603–1611. doi:10.1176/appi.ajp.161.9.1603

47. Tek C, Gold J, Blaxton T, Wilk C, McMahon RP, Buchanan RW. Visual Perceptual and Working Memory Impairments in Schizophrenia. Archives of General Psychiatry. 2002;59(2):146–153. doi:10.1001/archpsyc.59.2.146

48. Kumfor F, Liang CT, Hazelton JL, et al. Examining the presence and nature of delusions in Alzheimer’s disease and frontotemporal dementia syndromes. Int J Geriatr Psychiatry. 2022;37(3):10.1002/gps.5692. doi:10.1002/gps.5692

49. Drevets WC, Savitz J, Trimble M. The Subgenual Anterior Cingulate Cortex in Mood Disorders. CNS Spectr. 2008;13(8):663–681.

50. Riva-Posse P, Inman CS, Choi KS, et al. Autonomic arousal elicited by subcallosal cingulate stimulation is explained by white matter connectivity. Brain Stimulation. 2019;12(3):743–751. doi:10.1016/j.brs.2019.01.015

51. Lesage E, Stein EA. Networks Associated with Reward. In: Pfaff DW, Volkow ND, eds. Neuroscience in the 21st Century. Springer; 2016:1–27. doi:10.1007/978-1-4614-6434-1_134-1

52. Rolls ET. The cingulate cortex and limbic systems for emotion, action, and memory. Brain Struct Funct. 2019;224(9):3001–3018. doi:10.1007/s00429-019-01945-2

53. Alexander L, Clarke HF, Roberts AC. A Focus on the Functions of Area 25. Brain Sci. 2019;9(6):E129. doi:10.3390/brainsci9060129

54. Fornito A, Yücel M, Dean B, Wood SJ, Pantelis C. Anatomical Abnormalities of the Anterior Cingulate Cortex in Schizophrenia: Bridging the Gap Between Neuroimaging and Neuropathology. Schizophr Bull. 2009;35(5):973–993. doi:10.1093/schbul/sbn025

55. Fornito A, Yücel M, Wood SJ, et al. Anterior cingulate cortex abnormalities associated with a first psychotic episode in bipolar disorder. The British Journal of Psychiatry. 2009;194(5):426–433. doi:10.1192/bjp.bp.107.049205

56. Koo MS, Levitt JJ, Salisbury DF, Nakamura M, Shenton ME, McCarley RW. A Cross-Sectional and Longitudinal Magnetic Resonance Imaging Study of Cingulate Gyrus Gray Matter Volume Abnormalities in First-Episode Schizophrenia and First-Episode Affective Psychosis. Arch Gen Psychiatry. 2008;65(7):746–760. doi:10.1001/archpsyc.65.7.746

57. Brugger SP, Howes OD. Heterogeneity and Homogeneity of Regional Brain Structure in Schizophrenia: A Meta-analysis. JAMA Psychiatry. 2017;74(11):1104–1111. doi:10.1001/jamapsychiatry.2017.2663

58. Hosoda C, Tsujimoto S, Tatekawa M, Honda M, Osu R, Hanakawa T. Plastic frontal pole cortex structure related to individual persistence for goal achievement. Commun Biol. 2020;3(1):1–11. doi:10.1038/s42003-020-0930-4

59. Mansouri FA, Buckley MJ, Mahboubi M, Tanaka K. Behavioral consequences of selective damage to frontal pole and posterior cingulate cortices. Proceedings of the National Academy of Sciences. 2015;112(29):E3940–E3949. doi:10.1073/pnas.1422629112

60. Raju VB, Shukla A, Jacob A, et al. The frontal pole and cognitive insight in schizophrenia. Psychiatry Research: Neuroimaging. 2021;308:111236. doi:10.1016/j.pscychresns.2020.111236

61. Szczepanski SM, Knight RT. Insights into Human Behavior from Lesions to the Prefrontal Cortex. Neuron. 2014;83(5):1002–1018. doi:10.1016/j.neuron.2014.08.011

62. Masina F, Vallesi A, Di Rosa E, Semenzato L, Mapelli D. Possible Role of Dorsolateral Prefrontal Cortex in Error Awareness: Single-Pulse TMS Evidence. Frontiers in Neuroscience. 2018;12. Accessed June 8, 2022. https://www.frontiersin.org/article/10.3389/fnins.2018.00179

63. Jong H. Yoon MD, Michael J. Minzenberg MD, Stefan Ursu MD, et al. Association of Dorsolateral Prefrontal Cortex Dysfunction With Disrupted Coordinated Brain Activity in Schizophrenia: Relationship With Impaired Cognition, Behavioral Disorganization, and Global Function. American Journal of Psychiatry. Published online August 1, 2008. doi:10.1176/appi.ajp.2008.07060945

64. Sheth BR, Young R. Two Visual Pathways in Primates Based on Sampling of Space: Exploitation and Exploration of Visual Information. Frontiers in Integrative Neuroscience. 2016;10. Accessed June 15, 2022. https://www.frontiersin.org/article/10.3389/fnint.2016.00037

65. McLachlan E, Bousfield J, Howard R, Reeves S. Reduced parahippocampal volume and psychosis symptoms in Alzheimer’s disease. International Journal of Geriatric Psychiatry. 2018;33(2):389–395. doi:10.1002/gps.4757

66. Xu J, Wang J, Fan L, et al. Tractography-based Parcellation of the Human Middle Temporal Gyrus. Sci Rep. 2015;5:18883. doi:10.1038/srep18883

67. Adámek P, Langová V, Horáček J. Early-stage visual perception impairment in schizophrenia, bottom-up and back again. NPJ Schizophr. 2022;8(1):27. doi:10.1038/s41537-022-00237-9

68. Javitt DC, Freedman R. Sensory Processing Dysfunction in the Personal Experience and Neuronal Machinery of Schizophrenia. Am J Psychiatry. 2015;172(1):17–31. doi:10.1176/appi.ajp.2014.13121691

69. Chun CA, Brugger P, Kwapil TR. Aberrant Salience Across Levels of Processing in Positive and Negative Schizotypy. Frontiers in Psychology. 2019;10. Accessed June 21, 2022. https://www.frontiersin.org/article/10.3389/fpsyg.2019.02073

70. Kapur S. Psychosis as a State of Aberrant Salience: A Framework Linking Biology, Phenomenology, and Pharmacology in Schizophrenia. AJP. 2003;160(1):13–23. doi:10.1176/appi.ajp.160.1.13

71. Bassiony MM, Steinberg MS, Warren A, Rosenblatt A, Baker AS, Lyketsos CG. Delusions and hallucinations in Alzheimer’s disease: prevalence and clinical correlates. International Journal of Geriatric Psychiatry. 2000;15(2):99–107. doi:10.1002/(SICI)1099-1166(200002)15:2<99::AID-GPS82>3.0.CO;2-5

