## supplement Tables S2 and S3 for "Toward Localizing Psychosis in Pathologically Confirmed Neurodegenerative Disease"

### Supplementary Materials

**Table S1: Clinical Syndrome by Pathology**

| Pathology | | Psychosis | No Psychosis | Total |
|---|---|---|---|---|
| | **Clinical Syndrome** | | | |
| TDP | AD | 1 (33.3%) | 2 (66.7%) | 3 (3.9%) |
| | ALS | 1 (7.7%) | 12 (92.3%) | 13 (16.9%) |
| | ALS/bvFTD | 5 (29.4%) | 12 (70.6%) | 17 (22.1%) |
| | bvFTD | 13 (54.2%) | 11 (45.8%) | 24 (31.2%) |
| | CBS/PSP | 0 (0.0%) | 3 (100.0%) | 3 (3.9%) |
| | nfvPPA | 0 (0.0%) | 2 (100.0%) | 2 (2.6%) |
| | svPPA | 3 (20.0%) | 12 (80.0%) | 15 (19.5%) |
| Tau | AD | 1 (16.7%) | 5 (83.3%) | 6 (5.2%) |
| | ALS/bvFTD | 0 (0.0%) | 2 (100.0%) | 2 (1.7%) |
| | bvFTD | 10 (32.3%) | 21 (67.7%) | 31 (27.0%) |
| | CBS/PSP | 10 (17.5%) | 47 (82.5%) | 57 (49.6%) |
| | nfvPPA | 0 (0.0%) | 15 (100.0%) | 15 (13.0%) |
| | svPPA | 1 (25.0%) | 3 (75.0%) | 4 (3.5%) |
| AD | AD | 11 (22.0%) | 39 (78.0%) | 50 (79.4%) |
| | ALS/bvFTD | 0 (0.0%) | 1 (100.0%) | 1 (1.6%) |
| | bvFTD | 1 (33.3%) | 2 (66.7%) | 3 (4.8%) |
| | CBS/PSP | 2 (33.3%) | 4 (66.7%) | 6 (9.5%) |
| | DLB/PD | 1 (100.0%) | 0 (0.0%) | 1 (1.6%) |
| | nfvPPA | 0 (0.0%) | 2 (100.0%) | 2 (3.2%) |
| LBD/AD | AD | 7 (36.8%) | 12 (63.2%) | 19 (86.4%) |
| | CBS/PSP | 1 (100.0%) | 0 (0.0%) | 1 (4.5%) |
| | DLB/PD | 1 (50.0%) | 1 (50.0%) | 2 (9.1%) |

**Table S2: Copathologies by Primary Pathology Category**

| Psychosis | Primary Pathology Category | Total | ADNC | LBD | TDP43 | Tauopathy | AGD | HS | PLS | MND | ATAC |
|---|---|---|---|---|---|---|---|---|---|---|---|
| Absent | AD | 48 | 48 (100%) | 13 (27%) | 9 (19%) | 16 (33%) | 14 (29%) | 3 (6%) | 1 (2%) | 0 (0%) | 4 (8%) |
| | LBD_AD | 13 | 13 (100%) | 13 (100%) | 2 (15%) | 7 (54%) | 7 (54%) | 2 (15%) | 0 (0%) | 0 (0%) | 1 (8%) |
| | TDP | 54 | 42 (78%) | 4 (7%) | 54 (100%) | 20 (37%) | 12 (24%) | 3 (6%) | 0 (0%) | 27 (50%) | 0 (0%) |
| | tauopathy | 93 | 65 (70%) | 13 (14%) | 6 (7%) | 93 (100%) | 19 (21%) | 2 (2%) | 0 (0%) | 1 (1%) | 4 (4%) |
| | Other | 5 | 4 (80%) | 1 (20%) | 2 (40%) | 1 (20%) | 1 (20%) | 1 (20%) | 0 (0%) | 0 (0%) | 0 (0%) |
| Present | AD | 15 | 15 (100%) | 3 (20%) | 2 (13%) | 4 (27%) | 3 (20%) | 1 (7%) | 1 (7%) | 0 (0%) | 2 (13%) |
| | LBD_AD | 9 | 9 (100%) | 9 (100%) | 1 (11%) | 3 (33%) | 3 (33%) | 0 (0%) | 0 (0%) | 0 (0%) | 1 (11%) |
| | TDP | 23 | 13 (57%) | 4 (17%) | 23 (100%) | 13 (57%) | 9 (43%) | 1 (4%) | 1 (4%) | 11 (48%) | 1 (4%) |
| | tauopathy | 22 | 15 (68%) | 3 (14%) | 1 (5%) | 22 (100%) | 6 (27%) | 0 (0%) | 0 (0%) | 0 (0%) | 0 (0%) |
| | Other | 1 | 1 (100%) | 1 (100%) | 0 (0%) | 1 (100%) | 1 (100%) | 0 (0%) | 0 (0%) | 0 (0%) | 0 (0%) |

Table includes all study participants, organized by primary and secondary neuropathologies.

ADNC: Alzheimer Disease Neuropathological Changes are present (Montine et al. Acta Neuropathol. 2012)

LBD: Lewy Body Disease present

TDP-43: TDP-43 proteinopathy present

AGD: Argyrophilic Grain Disease

HS: Hippocampal sclerosis present

PLS: Primary Lateral Sclerosis

MND: Motor Neuron Disease

ATAC: Argyrophilc thorny astrocytes in clusters present

Note: There were several cases where information about specific copathologies was not available, which included 1 case for TDP43, 6 cases for AGD, and 3 cases for ATAC.

**Table S3: Lewy Body Disease Pathology Subcategories by Primary Pathology Category**

| Psychosis | Primary Pathology Category | LBD Total | Neocortical | Transitional | Brainstem only | Amygdala only | DMNV | Amygdala and DMNV | Nonspecific | Substantia Nigra |
|---|---|---|---|---|---|---|---|---|---|---|
| Absent | **AD** | 13 (27%) | 0 (0%) | 0 (0%) | 1 (2.1%) | 6 (12.5%) | 0 (0%) | 1 (2.1%) | 5 (10.4%) | 0 (0%) |
| | **LBD_AD** | 13 (100%) | 5 (38.5%) | 8 (61.5%) | 0 (0%) | 0 (0%) | 0 (0%) | 0 (0%) | 0 (0%) | 0 (0%) |
| | **Other** | 1 (20%) | 1 (20%) | 0 (0%) | 0 (0%) | 0 (0%) | 0 (0%) | 0 (0%) | 0 (0%) | 0 (0%) |
| | **TDP** | 4 (7%) | 0 (0%) | 1 (1.9%) | 1 (1.9%) | 2 (3.7%) | 0 (0%) | 0 (0%) | 0 (0%) | 0 (0%) |
| | **tauopathy** | 13 (14%) | 1 (1.1%) | 1 (1.1%) | 9 (9.7%) | 1 (1.1%) | 0 (0%) | 0 (0%) | 1 (1.1%) | 0 (0%) |
| Present | **AD** | 3 (20%) | 0 (0%) | 0 (0%) | 0 (0%) | 3 (20%) | 0 (0%) | 0 (0%) | 0 (0%) | 0 (0%) |
| | **LBD_AD** | 9 (100%) | 7 (77.8%) | 2 (22.2%) | 0 (0%) | 0 (0%) | 0 (0%) | 0 (0%) | 0 (0%) | 0 (0%) |
| | **Other** | 1 (100%) | 0 (0%) | 1 (100%) | 0 (0%) | 0 (0%) | 0 (0%) | 0 (0%) | 0 (0%) | 0 (0%) |
| | **TDP** | 4 (17%) | 0 (0%) | 1 (4.3%) | 1 (4.3%) | 1 (4.3%) | 1 (4.3%) | 0 (0%) | 0 (0%) | 0 (0%) |
| | **tauopathy** | 3 (14%) | 0 (0%) | 2 (9.1%) | 0 (0%) | 0 (0%) | 0 (0%) | 0 (0%) | 0 (0%) | 1 (4.5%) |
| **Total** | | 64 | 14 | 16 | 12 | 13 | 1 | 1 | 6 | 1 |

Table includes all individuals with Lewy body disease (N=64) as either a primary or copathology, with extent of synuclein infiltration; DMNV: Dorsal Motor Nucleus of Vagus

**Table S4:** Anatomical correlates of psychosis subtypes by pathology

| **Anatomical correlates of psychosis subtypes by pathology** | | | | | | | | | | | |
|---|---|---|---|---|---|---|---|---|---|---|---|
| | **MaxT by psychosis subtype** | | | | | **Right** | | | **Left** | | |
| **Anatomic Structure** | Any Psychosis | Any Delusions | Paranoid Delusions | Persecutory Delusions | Misidentification Delusions | x | y | z | x | y | z |
| **TDP Pathology (N=77)** | | | | | | | | | | | |
| ***Frontal Lobe*** | | | | | | | | | | | |
| Left superior frontal gyrus | | | | 3.96 | | | | | -6 | 61 | 21 |
| Right superior frontal gyrus | 4.19 | | | | | 24 | 31 | 46 | | | |
| **Tau Pathology (N=115)** | | | | | | | | | | | |
| ***Temporal Lobe*** | | | | | | | | | | | |
| **Right temporal pole** | **5.40** | 4.26 | 3.65 | 4.04 | | 33 | 22 | -30 | | | |
| Right entorhinal area | 4.23 | | | | | 30 | 6 | -19 | | | |
| Right inferior temporal gyrus/fusiform gyrus | | | | 4.26 | | 33 | -16 | -36 | | | |
| ***Frontal Lobe*** | | | | | | | | | | | |
| **Right superior frontal gyrus medial segment** | **5.05** | 4.20 | 4.18 | **4.81** | | 4 | 52 | 8 | | | |
| Left superior frontal gyrus medial segment | | | | 4.72 | | | | | -6 | 46 | 4 |
| Right medial frontal cortex | | | 4.27 | 4.34 | | 1 | 43 | -12 | | | |
| Right superior frontal gyrus | 4.53 | | | | | | | | | | |
| **Right frontal pole** | | | | **4.88** | | 19 | 63 | -9 | | | |
| **Right subcallosal area** | 4.19 | 4.39 | 4.26 | **5.15** | | 1 | 21 | -7 | | | |
| Right anterior cingulate gyrus (pregenual) | | | 4.33 | | | 2 | 43 | 1 | | | |
| Right anterior cingulate gyrus (anterior subgenuall) | | | | 4.66 | | 2 | 34 | -9 | | | |
| Right accumbens area | | | | 4.39 | | 6 | 10 | -9 | | | |
| Left accumbens area | | | | 4.34 | | | | | -6 | 10 | -7 |
| Right hippocampus | | | | 4.05 | | 24 | -9 | -18 | | | |
| **Left subcallosal area** | | 4.01 | 4.21 | **5.03** | | | | | -2 | 15 | -6 |
| **Left anterior cingulate gyrus (pregenual)** | | | 3.92 | **4.99** | | | | | -3 | 40 | -3 |

| | | | | | | | | | | | | |
|---|---|---|---|---|---|---|---|---|---|---|---|---|
| **Left anterior cingulate gyrus (pregenual)** | | | 3.86 | **4.86** | | | | | | -3 | 33 | -9 |
| **Left frontal pole** | | | 3.88 | **4.88** | | | | | | -12 | 66 | -4.5 |
| **Right gyrus rectus** | | | 3.81 | **4.78** | | | 4 | 49 | -22 | | | |
| Left gyrus rectus | 4.21 | | | 4.09 | | | | | | -2 | 47 | -22 |
| Left anterior orbital gyrus | | 4.18 | | 4.18 | | | | | | -30 | 58 | -8 |
| Left medial orbital gyrus | | | | 4.33 | | | | | | -13 | 19 | -22 |
| Right middle frontal gyrus | 3.97 | | | 3.91 | | | 44 | 49 | -5 | | | |
| Right posterior orbital gyrus | | | | 4.27 | | | 25 | 33 | -12 | | | |
| **AD Pathology (N=63)** | | | | | | | - | | | | | |
| ***Parietal/Occipital Lobe*** | | | | | | | | | | | | |
| **Right superior occipital gyrus** | **5.27** | **4.93** | 4.10 | | | | 25 | -75 | 46 | | | |
| Right parahippocampal gyrus | 4.36 | | | | | | 28 | -27 | -25 | | | |
| Right angular gyrus | 4.01 | | | | | | 51 | -45 | 22 | | | |
| Right middle occipital gyrus | | | | | | 4.57 | 39 | -78 | 33 | | | |

**Figure S1**: Areas of significant atrophy within pathology group by form of psychosis

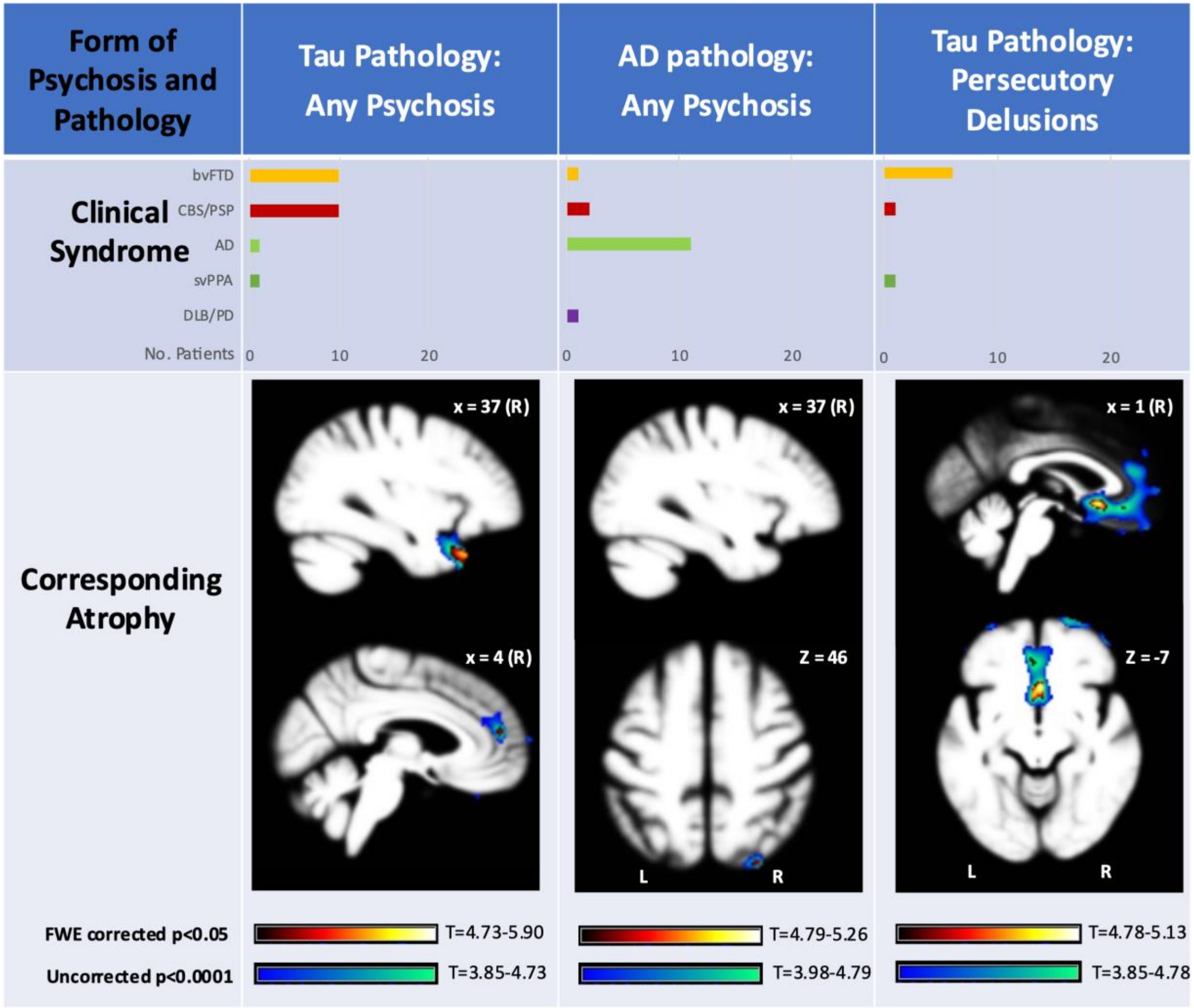
